# Inflammatory leptomeningeal cytokines mediate delayed COVID-19 encephalopathy

**DOI:** 10.1101/2020.09.15.20195511

**Authors:** Jan Remsik, Jessica A. Wilcox, N. Esther Babady, Tracy A. McMillen, Behroze A. Vachha, Neil A. Halpern, Vikram Dhawan, Marc Rosenblum, Christine A. Iacobuzio-Donahue, Edward K. Avila, Bianca Santomasso, Adrienne Boire

**Author notes:** Corresponding author: Adrienne Boire. Co-first authors: Jan Remsik and Jessica A. Wilcox.

## Abstract

SARS-CoV-2 infection induces a wide spectrum of neurologic dysfunction. Here we show that a particularly vulnerable population with neurologic manifestations of COVID-19 harbor an influx of inflammatory cytokines within the cerebrospinal fluid in the absence of viral neuro-invasion. The majority of these inflammatory mediators are driven by type 2 interferon and are known to induce neuronal injury in other disease models. Levels of matrix metalloproteinase-10 within the spinal fluid correlate with the degree of neurologic dysfunction. Furthermore, this neuroinflammatory process persists weeks following convalescence from the acute respiratory infection. These prolonged neurologic sequelae following a systemic cytokine release syndrome lead to long-term neurocognitive dysfunction with a wide range of phenotypes.

## Main text

Emerging hospital series demonstrate that acute respiratory infection with SARS-CoV-2 is frequently associated with neurologic dysfunction. Mild neurologic symptoms, including headaches, early anosmia and dysgusia, occur in a large portion of the infected population and typically resolve^1^. More serious complications, such as protracted delirium, seizures, and meningoencephalitis, appear to afflict more critically ill patients with hypoxic respiratory failure and may be devastating to higly susceptible individuals^2^. Cancer patients, in particular, are at heightened risk of severe infections from COVID-19 due to their baseline immunocompromised state and poor functional reserve^3^. The mechanism by which COVID-19 impacts the CNS is unclear, with hypotheses including direct viral neuroinvasion, neurologic toxicity from the systemic cytokine release syndrome, or a combination of both. Investigations to date lack consensus regarding neuroinvasion potential of COVID-19, with detectable SARS-CoV-2 virus present within the CSF in only a small number of patients with neurologic toxicity^2,4-6^. Small case reports of 1 to 3 patients have reported an elevation in pro-inflammatory cytokines, such as IL-6, CXCL10 (also known as IFN-induced protein-10), and CCL2, in the spinal fluid of acutely infected individuals^6-8^. No available literature has characterized the full extent and duration of the neuro-inflammatory response to COVID-19 in a large sample of patients, or compared the degree of neuroinflammation with the severity of neurologic dysfunction. Here we present complete clinical neurologic characterization of cancer patients with neurologic toxicity following SARS-CoV-2 infection correlated with biochemical analysis of CSF.

Between May and July 2020, we prospectively evaluated 18 cancer patients with confirmed SARS-CoV-2 respiratory infection who subsequently developed moderate to severe neurologic symptoms (**Table 1** and **Supplementary Table 1**). COVID-19 nasopharyngeal swab was positive in 16 patients; two additional patients were included on the basis of a positive serum COVID-19 antibody screen and recent respiratory illness consistent with COVID-19 (**Supplementary Table 2**). All patients presented with classic features of a COVID-19 respiratory infection including cough, dypsnea and fever; eleven patients experienced severe hypoxic respiratory failure and cytokine release syndrome requiring mechanical ventilatory support (**Supplementary Table 3)**. Patients displayed a wide range of neurologic manifestations, including prolonged hyperactive or hypoactive delirium (n = 10), limbic encephalitis (n = 4), refractory headaches (n = 2), rhombencephalitis (n = 1), and large-territory infarctions (n = 1). There was a delay of 19 days (median, range 0 – 77 days) between the onset of respiratory infection and clincal documentation of neurological symptoms.

**Table 1.** Pathologic findings in cancer patients with neurologic manifestations of COVID-19.

|  | Median or n | Range or % |
| --- | --- | --- |
| **Patient characteristics** |  |  |
| Active cancer or history of cancer | 18/18 | 100 |
| Time from respiratory illness to onset of neurologic symptoms (days) | 19.0 | 0 - 77 |
| Baseline Karnofsky performance score (KPS) | 80 | 60 - 100 |
| KPS at lumbar puncture | 50 | 20 - 70 |
| DRS at lumbar puncture | 13 | 1 - 26 |
| **Neurologic syndrome (n = 18)** |  |  |
| Prolonged delirium | 10/18 | 55.6% |
| Limbic encephalitis | 4/18 | 22.2% |
| Refractory headaches | 2/18 | 11.1% |
| Rhombencephalitis | 1/18 | 5.6% |
| Ischemic infarcts | 1/18 | 5.6% |
| **Electroencephalography (n = 14)** |  |  |
| Posterior dominant rhythm (PDR) | 7.5 | 5 - 9 |
| Absent PDR | 6/14 | 42.9% |
| Sedatives | 4/14 | 28.6% |
| EEG abnormalities: |  |  |
| Diffuse slowing | 12/14 | 85.7% |
| Focal slowing | 5/14 | 35.7% |
| Triphasic waves | 4/14 | 28.6% |
| Seizures | 1/14 | 10.0% |
| GRDA | 1/14 | 10.0% |
| **Neurologic imaging (MRI n = 14, CT n = 4)** |  |  |
| Increased T2-signal in focal cortical and white matter structures (MRI): | 3/14 | 21.4% |
| Limbic regions | 2/14 | 14.3% |
| Cerebellum | 1/14 | 7.1% |
| New diffuse microhemorrhages (MRI) | 1/14 | 7.1% |
| Large territory infarcts (MRI + CT) | 1/18 | 5.6% |
| Increased white matter disease from prior MRI (COVID-19 non-related) | 3/14 | 36.4% |
| Abnormal contrast enhancement, excluding brain metastasis (MRI) | 0/14 | 0.0% |

In addition to bedside neurologic examination, standard neurologic testing included neuroimaging in the form of brain MRI (n = 14) or head CT (n = 4) (**Table 1** and **Extended Data Fig. 1**). Three patients demonstrated encephalitic changes in the form of non-enhancing T2-hyperintense white matter and cortical changes afflicting the limbic or cerebellar structures. Large territory infarcts, diffuse microhemorrhages, and increased subcortical or periventricular white disease were also evident. Electroencephalography (**Table 1**) was completed in 14 patients. Diffuse bihemispheric slowing was the most common finding observed (n = 12), and was irrespective of the use of pharmacologic sedation. A single patient with limbic encephalitis developed temporal lobe seizures.

Thirteen patients underwent spinal fluid analysis to rule out alternative causes of meningoencephalitis, at a median of 57 days from the onset of respiratory symptoms (**Supplementary Table 4**). Notably, cell count, protein, and glucose levels were normal; only 2 patients had measurable pleocytosis due to metastatic disease to the CNS. CSF immune cell differentials were lymphocyte-predominant in 76.9% and monocyte-predominant in 23.1% of patients. Oligoclonal bands were detected in both the serum and CSF in 83% of tested patients, indicating systemic rather than intracerebral production of gammaglobulins. Only two patients had elevated intracranial pressure and abnormal protein levels, a sequela attributed to known brain metastases rather than COVID-19 neurologic toxicity.

To characterize COVID-19 neuroinvasion within this cancer population, we optimized a PCR-based assay for the CSF and used a commercially available ELISA to test for SARS-CoV-2 viral RNA and N and S structural proteins, respectively, in the CSF. No patients demonstrated detectable levels of viral PCR or structural proteins in the CSF, supporting prior reports^4,9,10^. We also tested the levels of ACE2, the SARS-CoV-2 receptor, within the CSF-brain and CSF-blood barriers in banked pre-pandemic cerebral tissues. We detected low levels of ACE2 in the intravascular monocytes and capillary endothelium within both the choroid plexus and leptomeninges, representing potential means of viral entry into the leptomeninges (**Extended Data Fig. 2)**. Anti-N COVID-19 IgG were detected in the CSF of only one patient with limbic encephalitis and temporal lobe seizures, indicating blood-brain barrier breakdown^11^ (**Extended Data Fig. 1** and **Supplementary Table 5**).

As direct viral invasion could not explain neurologic dysfunction, we then subjected the CSF from 10 patients to an extensive proteomics assay to elucidate cytokine patterns. These results were compared to CSF from three control cohorts from the pre-pandemic era. To control for alternative cancer-related causes of CNS inflammation, the first group consisted of patients matched for age, cancer type, and presence of brain metatases (**Supplementary Table 6**). To compare with other well-characterized neuro-inflammatory syndromes in cancer patients, the second group consisted of patients with CAR T cell-associated neurotoxicity, also known as ICANS^12^ (Immune Effector Cell Associated Neurotoxicity Syndrome), and the third group with autoimmune encephalitis (idiopathic n = 3 and suspected immune checkpoint inhibitor-associated n = 3)^13,14^. Correlative analysis across these 4 cohorts identified a significant accumulation of 12 inflammatory mediators in the CSF of COVID-19 patients, at levels approaching that of patients with severe CAR T-associated neurotoxicity (**Fig. 1A** and **Extended Data Fig. 3**). These included IL-6 and −8, IFN-γ, CXCL-1, −6, −9, −10, and −11, CCL-8 and −20, MMP-10 and 4E-BP1. Using an inflammatory signature comprising these 12 proteins, we also found a marked cumulative increase in inflammatory signalling relative to the non-COVID-19 cancer controls (p = 0.029, **Fig. 1B**). These 12 factors associate with cytokine and chemokine signaling, immune cell function, senescence, and neuroinflammation (**Extended Data Fig. 4**). We queried publicly available peripheral immune cell scRNA-sequencing data^15^ from patients with mild and severe COVID-19 infections to demonstrate that the majority of these factors appear to be absent peripherally and are likely isolated to the CNS compartment (**Extended Data Fig. 5)**, reminiscent of CAR associated neurotoxicity^13^.

**Fig. 1:**
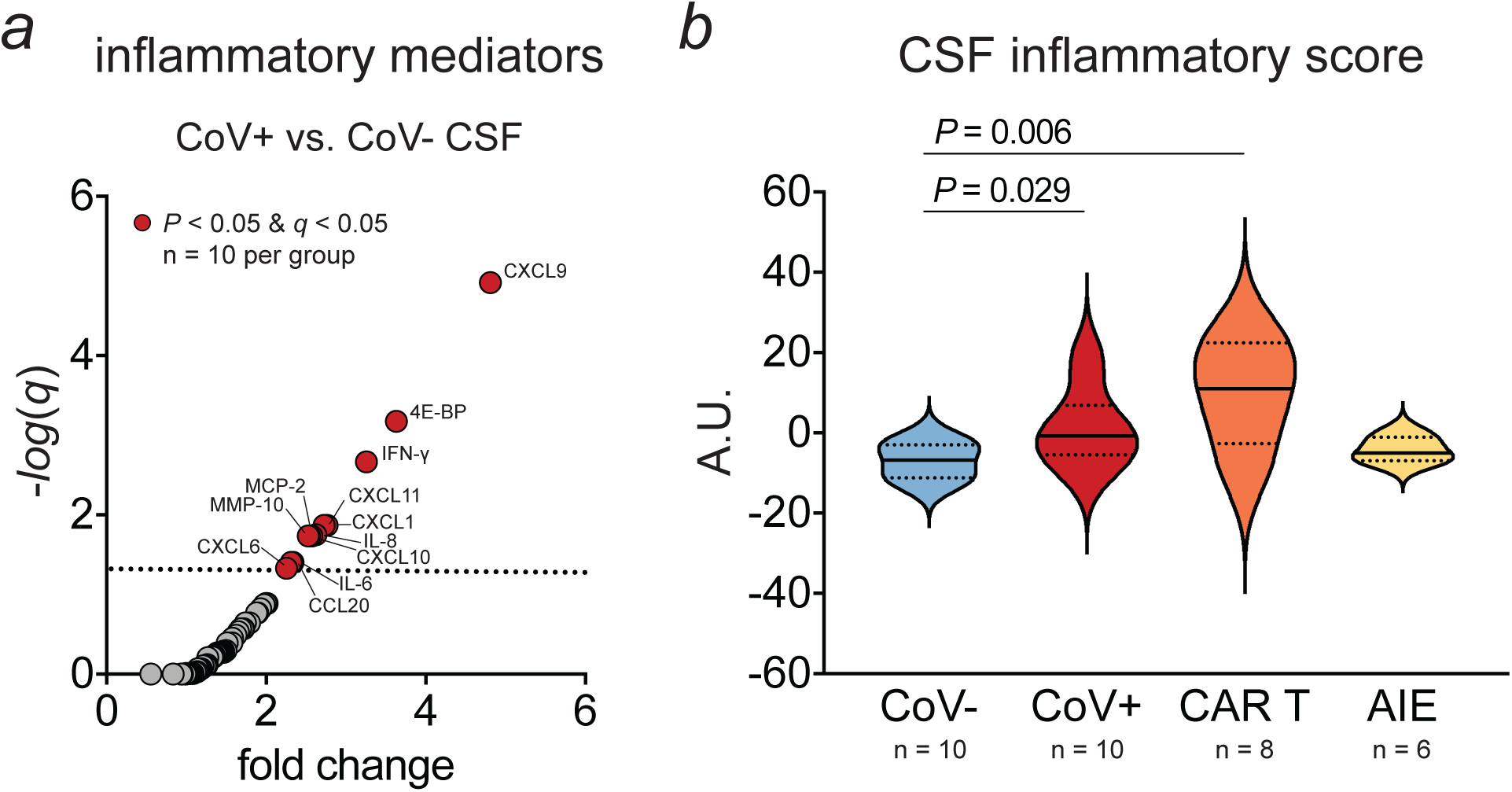
Accumulation of inflammatory mediators in the CSF of patients with neurologic manifestations of COVID-19. (a) Plot shows enrichment in 12 individual cytokines and chemokines in the CSF of cancer patients with COVID-19 (CoV+, n = 10) compared to their cancer-matched controls without COVID-19 (CoV-, n = 10; multiple t tests), as determined with targeted proteomic analysis. (b) Violin plots of a CSF inflammatory score incorporating these 12 mediators demonstrates statistically significant elevation of this neuroinflammatory signature in patients with COVID-19-related neurologic symptoms. CAR T cell neurotoxicity (CAR T) and autoimmune encephalitis (AIE) cohorts are shown as additional controls to represent other well-known neuroinflammatory syndromes (Full line = median, dotted line = quartiles; Whitney-Mann U test *P* reported within the plot; ANOVA Kruskal-Wallis test *P* = 0.015).

Moreover, we found that the CSF levels of MMP-10 in COVID-19 positive patients and controls correlated with clinical measures of neurologic dysfunction, by the Karnofsky performace status (KPS) and Disability Rating Scale^16^ (DRS) (**Extended Data Fig. 6**). This is consistent with other reports correlating MMP-10 levels with neurodegeneration, suggesting neuronal damage^17^. The specific mechanism through which COVID-19 induces this array of inflammatory molecules warrants further investigation, as does the extent of resulting neuronal damage.

In our COVID-19 positive patient cohort, a striking increase in IFN-γ and its downstream effectors, CXCL-9, −10, and −11, is demonstrated in the CSF nearly 2 months following onset of SARS-CoV-2 infection. As the major mediator against viral defences in both intracranial and systemic processes, the upregulation of this pathway is not unexpected. However, IFN-γ demonstrates opposing roles in other neuroinflammatory conditions, regulating both pro-inflammatory and neuroprotectant properties in oligodendroglial cells, microglia, and astrocytes^18^. Elevated levels of intracranial CXCL chemokines have been described in a wide array of infectious and non-infectious CNS pathologies.The subsequent intracerebral accumulation of monocyte-macrophages, T lymphocytes, and chemokines downstream of IFN-γ propogates further neurologic injury^19-22^, mirroring a pattern of lymphocytic infiltration identified in the brains of COVID-19 patients^23^. Similarly, a report following the 2003 SARS outbreak revealed that systemic response during an acute respiratory infection induced expression of interferon downstream effectors CXCL9 and −10 in glial cells, further propogating the destructive process^22^.

Our findings have potentially important diagnostic and treatment implications. A prolonged encephalopathy of unknown etiology accompanies critical infections with COVID-19, and studies to date have not identified any obvious meningitic, ischemic, or ictal cause. This is the first series to demonstrate an increase in pro-inflammatory CSF cytokines in such patients in the weeks following respiratory illness. Similar to other encephalidities, these cytokines may result in neuronal damage in the absence of obvious radiographic abnormalities.

Dexamethasone, a potent steroid and immunosuppressant, is the only intervention that has been shown in large-scale studies to reduce the incidence of death in hospitalized patients with severe COVID-19 infections^24^. The impact of dexamethasone on COVID-19 related neurologic toxicity, specifically, has not been defined. Our data demonstrate that the neurologic toxicity associated with COVID-19 is biochemically similar to CAR T cell neurotoxicity. Clinical rubrics for the grading and management of CAR T cell neurotoxicity support both early diagnosis and use of anti-inflammatory agents including dexamethasone^25-27^.

In conclusion, our data indicates that neurologic syndromes associated with moderate and severe COVID-19 infections are associated with a wide range of intracranial inflammatory cytokines, a finding that may persist for weeks to months following convalescence of the respiratory syndrome. The global accumulation of inflammatory mediators in CSF and inability to detect COVID-19 in the CSF suggests that the identification of a single, universal COVID-19 CSF biomarker is unlikely to be successful. However, the correlation of MMP-10 with neurologic dysfunction marks a potential prognostic biomarker for future study. Moreover, our findings further justify the investigation of early neurologic assessments, such those used for monitoring ICANS^12^, and use of anti-inflammatory therapies in patients with severe or prolonged COVID-19 disease.

## Abbreviations

ACE2: angiotensin-converting enzyme 2
AIE: autoimmune encephalitis
CAR T: chimeric antigen receptor T cell
CCL: C-C motif chemokine ligand
CRP: C-reactive protein
CSF: cerebrospinal fluid
CXCL: C-X-C motif chemokine ligand
DRS: disability rating scale
EEG: electroencephalography
ELISA: enzyme-linked immunosorbent assay
GRDA: generalized rhythmic delta activity
ICAN: Immune Effector Cell-Associated Neurotoxicity Syndrome
IFN: interferon
IL: interleukin
KPS: Karnofsky performance status
LP: lumbar puncture
MRI: magnetic resonance imaging
MMP-10: matrix metallopeptidase- 10
MCP2: monocyte chemoattractant protein-2
PCR: polymerase chain reaction
PDR: posterior dominant rhythm.

## Methods

### Ethical statement

This study was conducted according to the principles expressed in the Declaration of Helsinki. Ethical approval for COVID-19-relared research was obtained from MSKCC Institutional Review Board (IRB) under protocol #20-006. Collection and use of clinical samples were approved by MSKCC IRB #06-107, #12-245, #13-039, and #18-505. Only clinical samples collected in excess of those needed for diagnostic and therapeutic procedures were used for research. All participants provided written informed consent for sample and clinical data collection and subsequent analyses.

### Clinical sample collection

Eighteen cancer patients with recent or history of recent respiratory illness or COVID-19 with moderate to severe neurologic symptoms were evaluated by the MSK Neurology Consult Service between May and July 2020. All patients tested positive for the presence of COVID-19 viral RNA in nasopharyngeal swab and/or anti-COVID-19 antibodies in serum or plasma as described below, using CDC and FDA approved methods. The neurological symptoms are described in Table 1 and the detailed demographic characteristics are listed in Supplementary Table 1. Cerebrospinal fluid in excess of that needed for diagnostic purposes was collected via bedside or fluoroscopically-guided lumbar puncture. At the time of spinal fluid collection, venous blood was collected into a tube containing sodium citrate, or sodium or lithium heparin. CSF samples were kept on ice and processed within two hours from collection. Anti-coagulated blood and CSF were transferred into 15 mL conical tubes and spun at 800g for 10 minutes. Plasma and cell-free CSF was aliquoted and stored at −80°C. Processing and experimentation with non-inactivated body fluids were performed under Bsl2+ containment.

### SARS-CoV-2 RNA and protein detection

Total nucleic acids was extracted from CSF (200 µL) on the MagNA Pure compact (Roche Molecular Diagnostics) and eluted in a final volume of 100 µL. Real-time reverse transcriptase PCR was performed on an ABI 7500 Fast (Applied Biosystems) using 5 µL of extracted nucleic acids and primers and probes targeting two regions of the nucleocapsid gene (N1 and N2) and the human RNase P gene as in internal control as previously described for respiratory samples^28^. Results were valid only if the RNase P gene was detected in the sample. Samples were considered positive if both analytic targets N1 and N2 were detected and negative if both targets were not detected. N and S (S2 subunit) SARS-CoV-2 structural proteins were detected using commercially available ELISA kits (ELV-COVID19N-1 and ELV-COVID19S2-1, RayBiotech), as recommended by the manufacturer. All ELISA samples were run in technical replicates.

### Magnetic resonance imaging

Imaging studies were conducted either on 1.5 or 3 Tesla MRI. Images were obtained as part of the patient’s routine standard of care imaging, which did not allow standardization of sequences. The most frequently sequences performed were diffusion-weighted imaging (DWI), susceptibility-weighted angiography (SWAN), 2D fluid-attenuated inversion recovery (FLAIR), T2-weighted fast spin-echo and T1-weighted fast spin-echo MRI before and after administration of gadolinium-based contrast agent. MRI parameters of the most commonly used sequences in this study are tabulated in Supplementary Table 7.

### Electroencephalography

Long-term and routine electroencephalography monitoring with video was performed using a Natus 32 channel computerized EEG system with digital analysis of video-EEG for spike detection and analysis, computerized automated seizure detection algorithms, and patient “event marker.” Electrode set up of 8 channels or greater was performed by the EEG technologist. Standard 10-20 system montages were employed for EEG review.

### Clinical laboratory analyses

Levels of CSF protein and glucose and serum D-dimer, ferritin, CRP and IL-6 were determined per standard laboratory methods using FDA-approved, in vitro diagnostics (IVD) kits.

### Detection of anti-SARS-CoV-2 immunoglobulins

Clinical IgG test against SARS-CoV-2 was performed using FDA EUA kit from Abbott (6R86-20). Experimental IgG tests against SARS-CoV-2 N and S1RBD proteins were detected in plasma and CSF using quantitative ELISA kits (IEQ-CoVN-IgG1 and IEQ-CoVS1RBD-IgG1, RayBiotech). Samples were analyzed as recommended by manufacturer, except that the plasma was diluted 1,500x and CSF 750x in 1x sample buffer. IgM and IgA against SARS-CoV-2 N protein were detected in plasma and CSF using semi-quantitative ELISA kits (IE-CoVN-IgM-1 and IE-CoVN-IgA-1, RayBiotech), as recommended by the manufacturer. All samples were run in technical replicates. These kits were for research use only and did not have FDA approval at the time of initial submission.

### ACE2 immunohistochemistry

Human autopsy tissue was collected under MSKCC IRB #18-065 and #18-292 from patients that provided written informed consent. Tissue was de-paraffinized, antigens were retrieved, and the procedure was performed essentially as described in ^29^. Primary anti-ACE-2 antibody (AF933, R&D) was used as recommended by the manufacturer, followed by the incubation with HRP-conjugated anti-goat secondary antibody (Immpress HRP Anti-Goat IgG, MP-7405, Vector Laboratories) and subsequently DAB EqV (SK-4103, Vector Laboratories). Nuclei were counterstained with hematoxylin (S3309, Dako). Stained, dehydrated slides were mounted in Vectamount (H-5000, Vector Laboratories), dried and scanned with Mirax (Zeiss).

### CSF proteomics and data analysis

CSF collected from patients with neurologic complications of COVID-19 via lumbar puncture was processed within two hours post collection, as described above, aliquoted and stored at −80°C. Retrospectively collected samples from primary and metastatic tumor-matched patients without COVID-19, patients with severe CAR T neurotoxicity (grade 3-4) and patients with autoimmune encephalitis were obtained from MSK Brain Tumor Center CSF Bank. Control patient cohort was selected from a random pool of potential matches. Samples were slowly thawed on ice and inactivated for COVID-19 as follows: 45 μL of CSF was mixed with 5 μL of 10% Triton X-100 (Sigma, T8787) in saline and incubated at room temperature for two hours. Samples were then dispensed in randomized fashion into 96-well PCR plate and stored at −80°C until further analysis. Relative levels of 92 inflammatory proteins were detected using proximity extension assay (Olink Target 96 Inflammation, Olink). Protein abundance values are shown in NPX units (*log2* scale). Analytical measuring range for each protein is available online (www.olink.com) or from the corresponding author upon request.

### Calculations of composite signature and computational analyses

Inflammatory signature was constructed as follows: z-score for each of the twelve analytes in this dataset was computed for all patients, the sum of all z-scores for a patient then represented an inflammatory score plotted in Figure 1. Pathway analysis was performed with Reactome (www.reactome.org) and IPA (Qiagen). Single-cell RNA-seq data from peripheral immune cells in patients with mild and severe COVID-19 were published in^15^ and accessed through FASTGenomics with implemented ScanPy and cellxgene (BD Rhapsody cohort Fresh PBMC & whole blood, Figure 7 in the original publication, accession SchulteSchrepping_2020_COVID19_Rhapsody_fresh PBMC-WB).

### Statistics

Sample size was not pre-determined and no patients, data points or samples were excluded. Differences in inflammatory protein abundance between COVID-19 positive subjects and control cohort were determined using multiple t tests (unpaired, two-tailed) with Benjamini and Hochberg correction for FDR. Proteins with *P* and q values lower than 0.05 were considered significant. Differences in inflammatory score between patient cohorts were determined with Mann-Whitney U test (unpaired, two-tailed). Inflammatory score was computed as described above. Number of replicates is stated in corresponding figure legends. For correlations, both Person’s and Spearman’s R is reported. Data used to generate figures in this study were submitted as “Source Data” tables. Statistical analyses were conducted in Prism (v8, GraphPad).

### Data availability

All data shown in the manuscript are available upon reasonable request from the corresponding author.

## Acknowledgements

We are deeply grateful to the patients and families who donated clinical samples and contributed to this research. We would like to thank Anne Reiner from MSKCC Epidemiology & Biostatistics for reviewing the statistical approaches in this study. This work was supported by NCI P30 CA008748 (Cancer Center Support Grant); and The Pew Charitable Trusts GC241069, The Damon Runyon Cancer Research Foundation GC240764, and The Pershing Square Sohn Cancer Research Alliance GC239280 (all to A.B.). J.R. is supported by The American Brain Tumor Association Basic Research Fellowship, The Terri Brodeur Breast Cancer Foundation Fellowship, and The Druckenmiller Center for Lung Cancer Research.

## Author contributions

A.B, B.S., J.A.W. and J.R. conceived the study. J.A.W. performed neurologic examinations, clinical annotations, and collected blood and CSF. J.R. performed the experiments, and analyzed and curated the data. N.A.H. and V.D. referred patients to neurology consult service. N.E.B and T.A.M developed and performed the CSF COVID-19 RNA PCR method. B.A.V. reviewed neuroimaging data. M.R. reviewed histological staining. C.I.-D. provided autopsy samples. E.K.A. reviewed EEG recordings. B.S. provided CSF from retrospective CAR T and AIE cohorts. J.R., J.A.W. and A.B. wrote the manuscript. A.B. supervised the study. All authors reviewed and approved the manuscript.

## Supplementary Information

**Extended Data Fig. 1:**
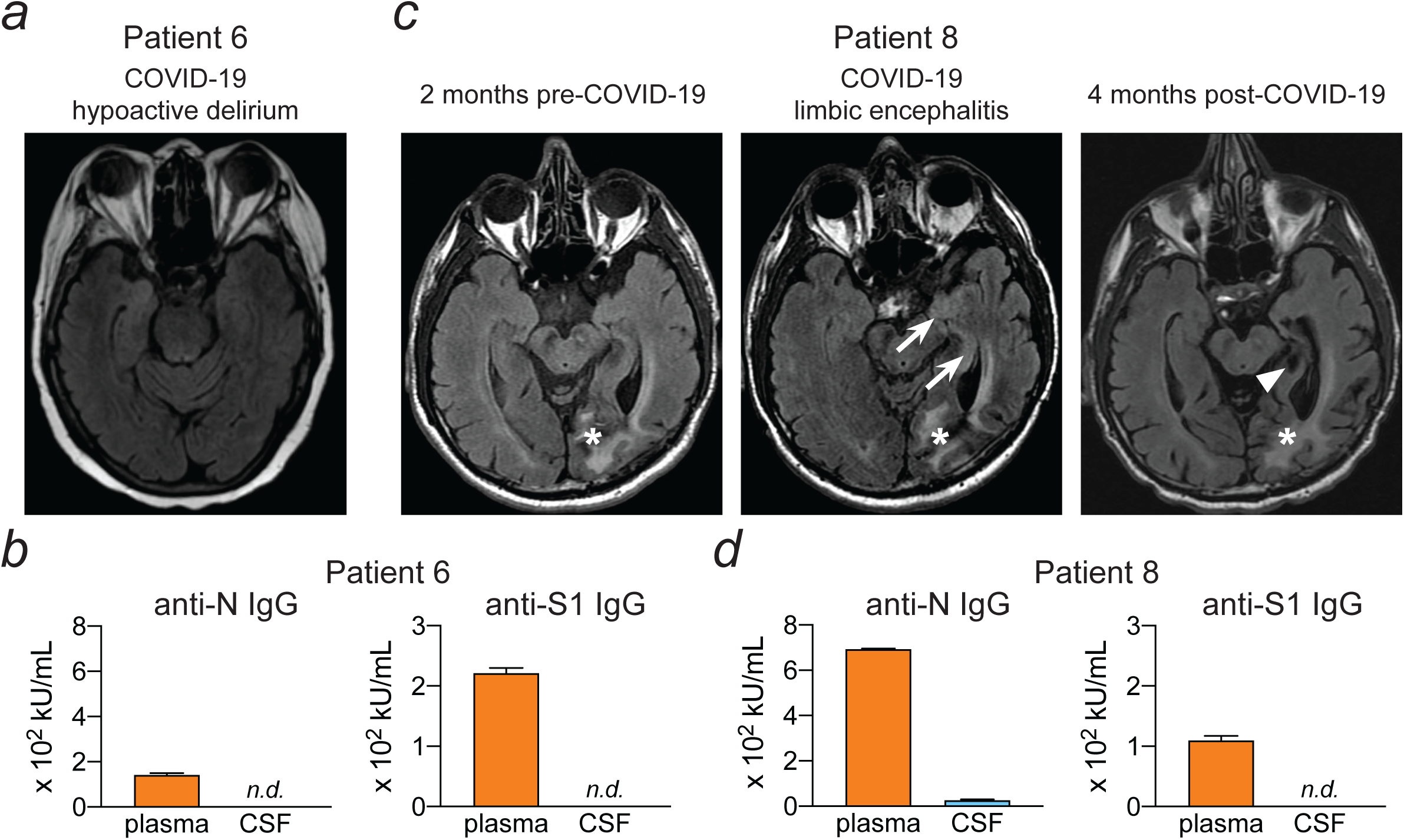
Radiographic findings and CSF COVID-19 antibodies in patients with different neurologic manifestations of COVID-19. Left: Patient 6 is a 71-year-old woman with NSCLC and prolonged hypoactive delirium following COVID-19 hypoxic respiratory failure. (a) Axial FLAIR sequences show no abnormalities. (b) Anti-N Covid-19 IgG is detectable in the plasma, but not in the CSF. Right: Patient 8 is a 61-year old man with NSCLC and COVID-19 limbic encephalitis. (c) Axial FLAIR images obtained 2 months prior to COVID-19 infection show baseline encephalomalacic post-operative changes (arrowheads, left). At the time of acute COVID-19 infection manifesting with refractory left temporal lobe seizures, axial FLAIR images demonstrate new T2 signal abnormality within the left medial temporal lobe (arrows, middle). Following resolution of COVID-19 limbic encephalitis with high-dose corticosteroids and antiepileptics, axial FLAIR images reveal improvement in medial temporal FLAIR abnormality, however new left hippocampal and parahippocampal gyrus atrophy (arrowhead, right). (d) Detectable levels of anti-N COVID-19 IgG are demonstrated in both the plasma and CSF of patient 8. n.d. = not detected.

**Extended Data Fig. 2:**
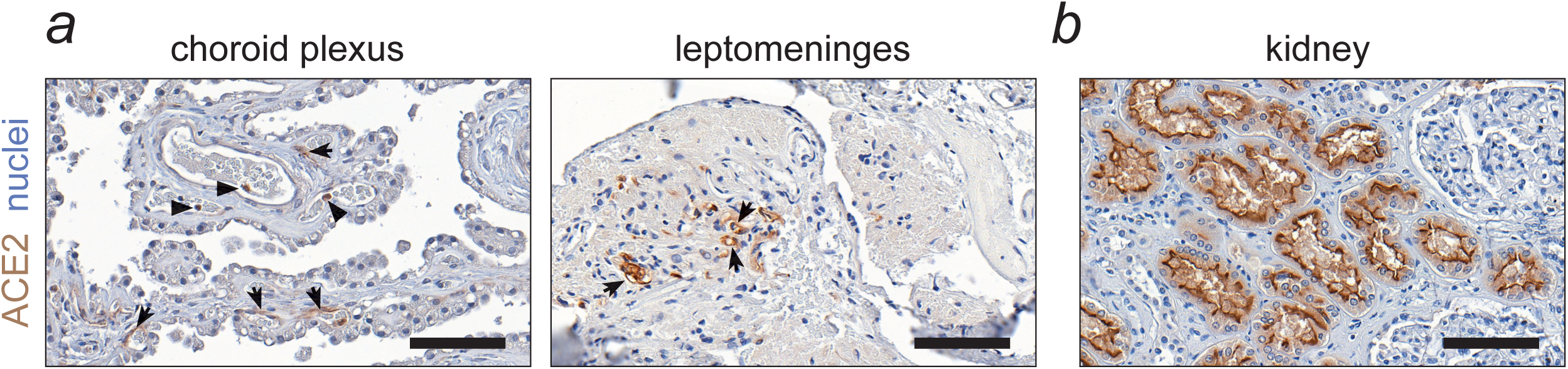
Expression of COVID-19 receptor ACE2 at CSF interfaces. (a) Representative images of human choroid plexus and leptomeninges (n = 3 per tissue) stained for ACE2. Note the moderate signal in the capillary endothelium (arrows) and intravascular monocytes (arrowheads). (b) Human kidney tubular epithelium, showing staining on the apical membrane, was used as a positive control (scale = 100 µm).

**Extended Data Fig. 3:**
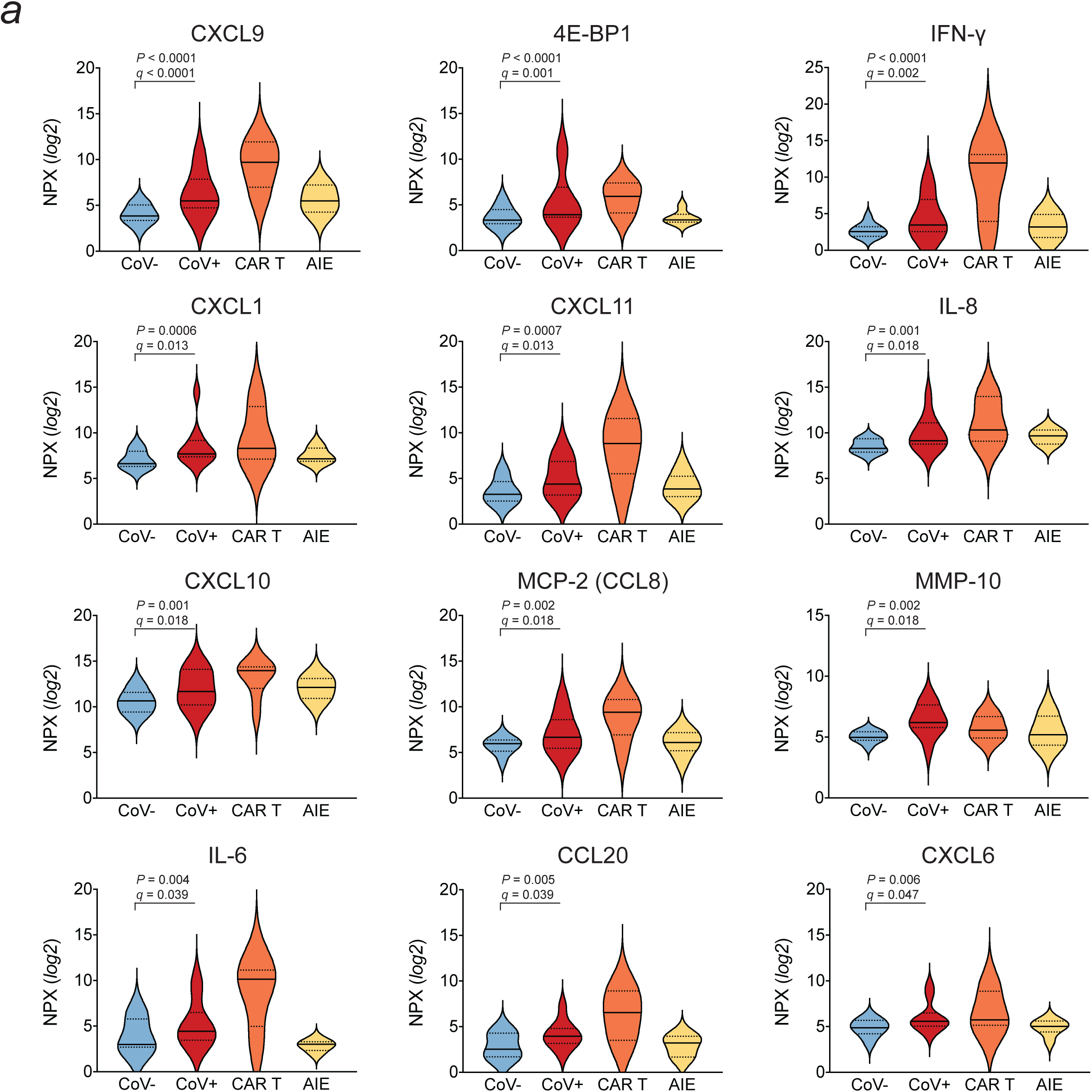
Differentially abundant inflammation mediators in the CSF of patients with neurologic consequences of COVID-19. (a) Violin plots show relative levels of 12 inflammatory mediators that are enriched in the CSF of patients manifesting with neurologic symptoms after COVID-19 infection (n = 10 per group, multiple t tests). Levels in the CSF from patients with CAR T cell neurotoxicity (CAR T; n = 8) and autoimmune encephalitis (AIE, n = 6) control groups are shown to represent other well-known neuroinflammatory syndromes. Full line = median, dotted line = quartiles.

**Extended Data Fig. 4.**
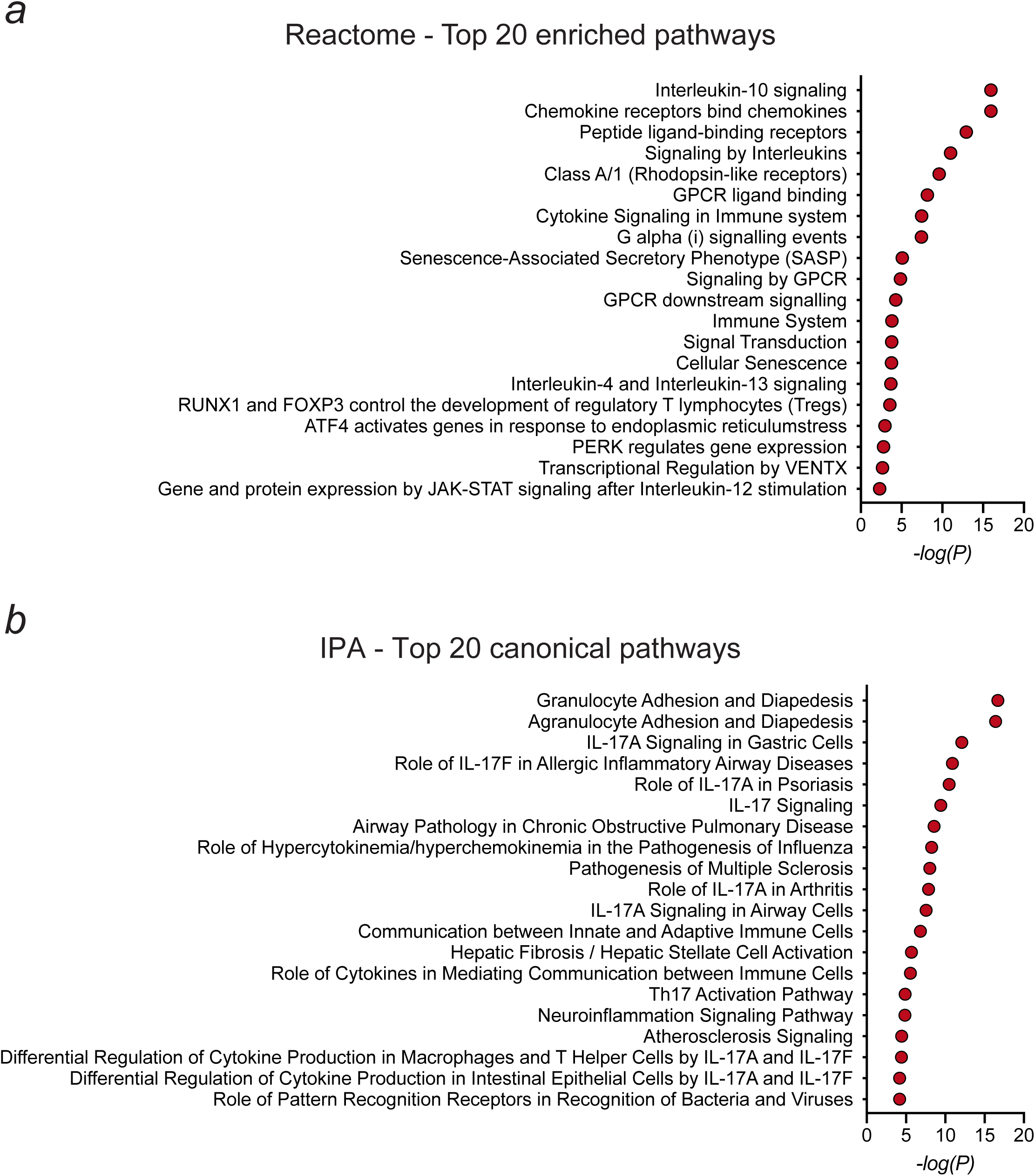
Pathway analysis of differentially abundant inflammatory mediators in the CSF of patients with neurologic symptoms of COVID-19. Plots show the top 20 pathways related to the 12 inflammatory mediators enriched in the CSF of COVID-19 patients, as analyzed with Reactome (a) and IPA (b).

**Extended Data Fig. 5:**
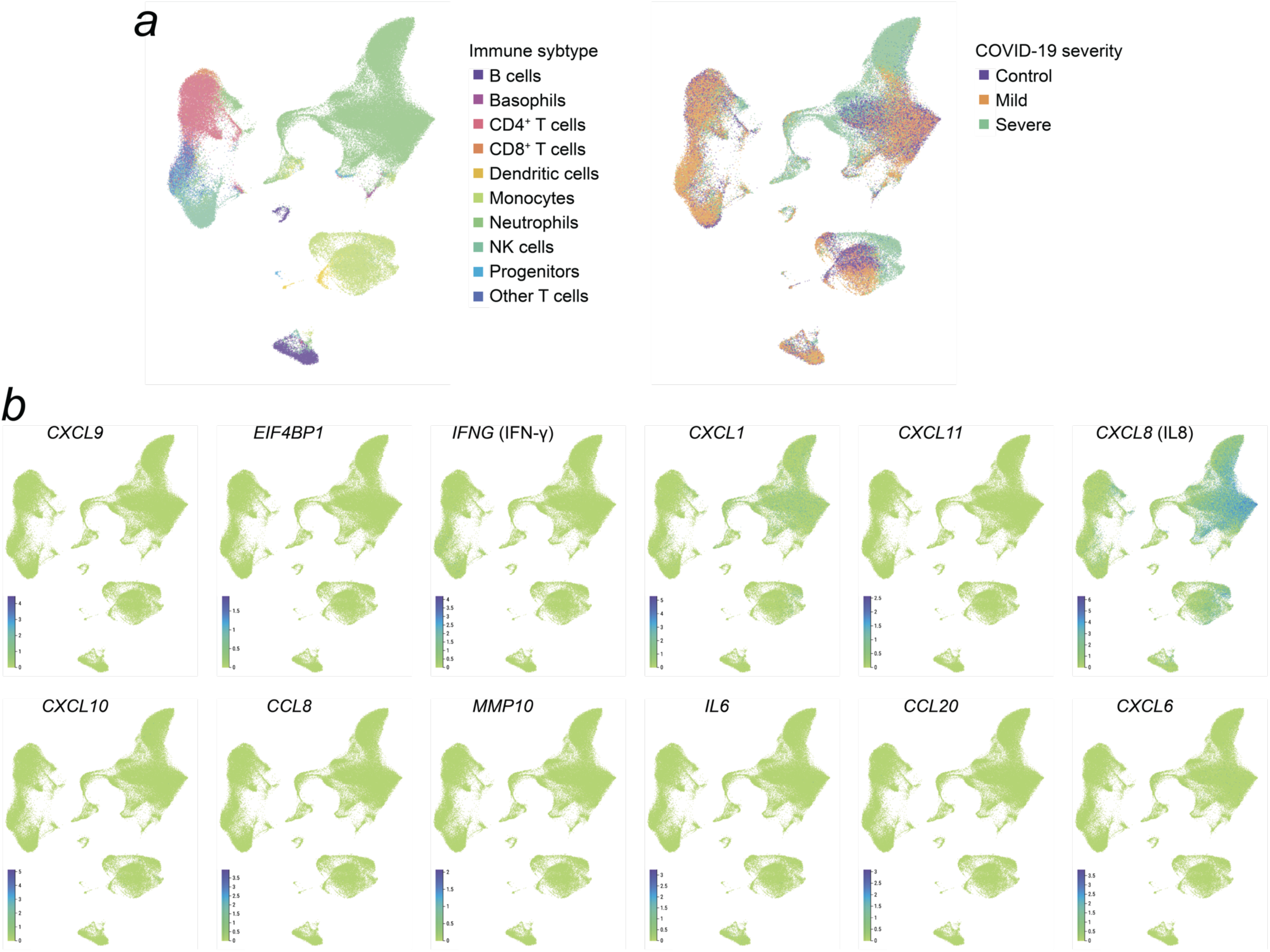
Expression levels of 12 differentially abundant inflammatory mediators in peripheral immune cells in patients with mild and severe COVID-19 infection. (a) UMAP plots show annotation of major immune cell clusters and distribution of cells based of COVID-19 infection severity (Fresh PBMC & whole blood cohort from Schulte-Schrepping et al., 2020). (b) Expression level projection of 12 differentially abundant CSF factors at single-cell resolution in the peripheral blood of patients with mild and severe COVID-19 infection (k= 122,954 cells; n = 17 control subjects, n = 6 patients with mild and n = 12 patients with severe COVID-19 infection). For details, please refer to the original publication.

**Extended Data Fig. 6:**
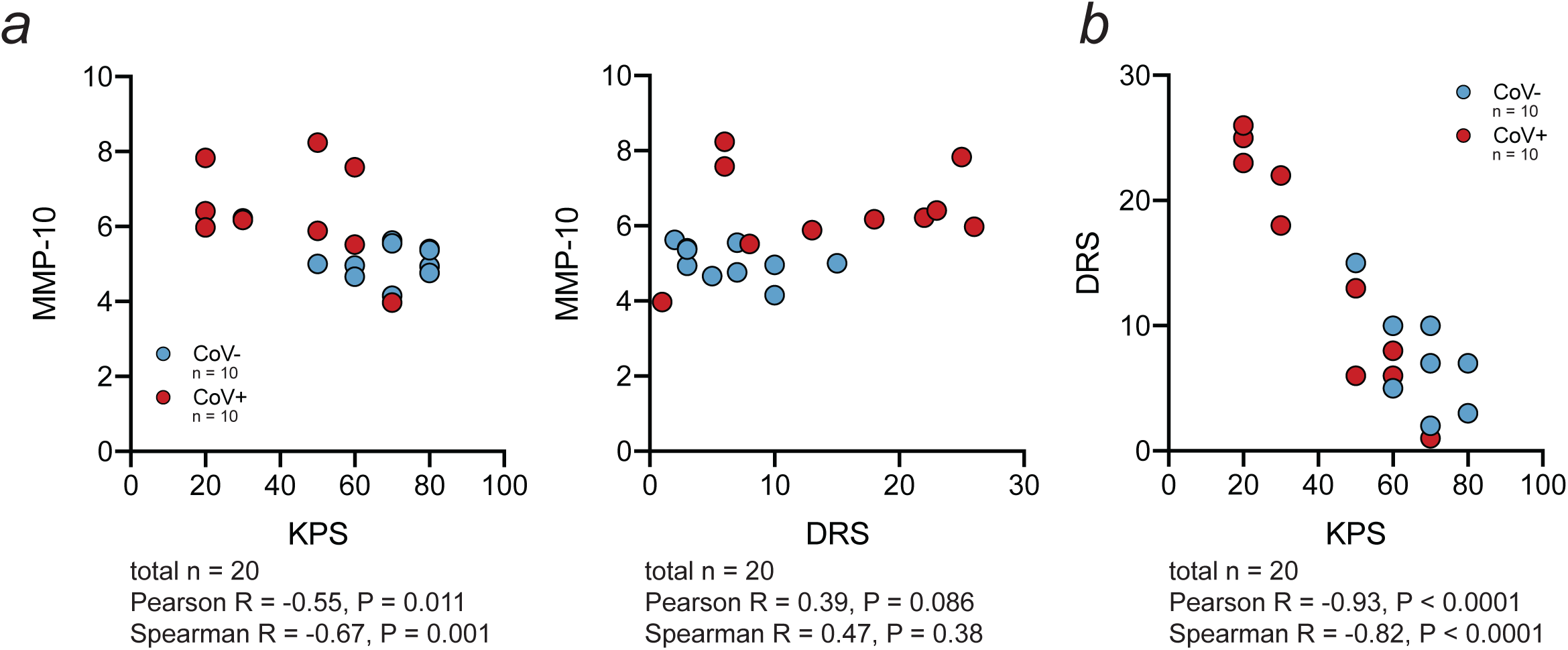
Levels of CSF MMP-10 correlate with clinical performance status and neurocognitive dysfunction in both control and COVID-19 patients. (a) Plots show positive correlation of MMP-10 with the degree of global (KPS) and neurocognitive (DRS) impairment in CoV- and CoV+ patients. (b) Plot confirming that KPS and DRS scores are inversely related, n = 10 per group. Please note that by scoring convention, KPS values (0 to 100) decrease and DRS values (0 to 29) increase with the degree of functional impairment.

**Supplementary Table 1.** Demographics and comorbidities of patients with neurological complications of COVID-19.

|  | n or value | Range or % |
| --- | --- | --- |
| **Age (mean)** | 64 | 41 - 80 |
| ≥ 65 | 10/18 | 55.6% |
| < 65 | 8/18 | 44.4% |
| **Sex** |  |  |
| Female | 9/18 | 50.0% |
| Male | 9/18 | 50.0% |
| **Medical Comorbidities** | 15/18 | 83.3% |
| Hypertension | 10/18 | 55.6% |
| Former smoker | 8/18 | 44.4% |
| Hyperlipidemia | 6/18 | 33.3% |
| Diabetes mellitus | 5/18 | 27.8% |
| Prior stroke | 1/18 | 5.6% |
| **Active Primary & Metastatic Cancer** | 17/18* | 94.4% |
| Lung | 6/18 | 33.3% |
| Lymphoma/Leukemia | 4/18 | 22.2% |
| Thyroid | 2/18 | 11.1% |
| Melanoma | 2/18 | 11.1% |
| Breast | 1/18 | 5.6% |
| Colorectal | 1/18 | 5.6% |
| Pancreatic | 1/18 | 5.6% |
| Ovarian | 1/18 | 5.6% |
| Brain metastases | 6/18 | 33.3% |
| Prior CNS-directed radiation | 5/18 | 27.8% |
| Cancer treatment within 30 days | 13/18 | 72.2% |
| *one patient did not have active cancer at the time of admission |  |  |
| **Baseline KPS** | 85 | 60 - 100 |

**Supplementary Table 2.** Characteristics of systemic COVID-19 infection in this patient cohort.

|  | n | % |
| --- | --- | --- |
| **COVID-19 at the admission to MSK** |  |  |
| Positive nasopharyngeal PCR | 16/18 | 88.9% |
| Prior respiratory illness with serum IgG | 2/18 | 11.1% |
| **COVID-19 systemic features** |  |  |
| Respiratory illness (cough, dyspnea, hypoxia) | 16/18 | 88.9% |
| Fever (>38.3°C) | 15/18 | 83.3% |
| ARDS | 11/18 | 61.1% |
| **Treatments** |  |  |
| Corticosteroids | 11/16 | 66.8% |
| Hydroxychloroquine | 10/16 | 62.5% |
| Remdesivir | 7/16 | 43.8% |
| Convalescent plasma | 4/16 | 25.0% |
| Tocilizumab | 3/16 | 18.8% |

**Supplementary Table 3.** Cerebrospinal fluid studies in this patient cohort.

|  | Median, mean or n | Range or % |
| --- | --- | --- |
| **Clinical Characteristics (n = 13)** |  |  |
| Time from onset of respiratory symptoms to LP (days) | 57 | 5 - 142 |
| KPS at LP | 50 | 20 - 70 |
| DRS at LP | 13 | 1 - 26 |
| **Cell counts, chemistry, and cytology (n = 13)** |  |  |
| Opening pressure (cm H<sub>2</sub>O) | 18 | 13-55 |
| Nucleated cell count (cells per microliter) | 1 | <1 - 147 |
| Lymphocyte-predominant | 10/12 | 76.9% |
| Monocyte-predominant | 3/12 | 23.1% |
| Neutrophil-predominant | 0/12 | 0.0% |
| Red blood cell count (cells per microliter) | 13 | <1 - 139 |
| Protein (mg/dL) | 32 | 19 - 218 |
| Glucose (mg/dL) | 37 | 21 - 100 |
| Positive cytology | 1/13 | 7.7% |
| **Oligoclonal bands (n = 12)** |  |  |
| No bands | 2/12 | 16.7% |
| 1-4 bands | 4/12 | 33.3% |
| ≥5 bands | 6/12 | 50.0% |
| **Immunoglobulin levels in CSF (n = 10)** |  |  |
| Normal | 7/10 | 70.0% |
| Elevated | 1/10 | 10.0% |
| **Detection of COVID-19 in the CSF (n = 13)** |  |  |
| Positive CSF PCR | 0/13 | 0.0% |
| N Nucleocapsid Protein ELISA (+) | 0/9 | 0.0% |
| S1 RBD Spike Protein ELISA (+) | 0/9 | 0.0% |

**Supplementary Table 4.** Laboratory serum studies confirming systemic inflammation in this patient cohort.

| **Laboratory serum studies (n = 10)** | **Median** | **Range** | **Reference** |
| --- | --- | --- | --- |
| D-dimer (peak; $\mu\text{g/mL}$) | 3.61 | 0.57 - 19.85 | 0.0 - 0.05 |
| Ferritin (peak; $\text{ng/mL}$) | 726 | 222 - 2598 | F 6 - 200, M 22 – 415* |
| CRP (peak; $\text{mg/dL}$) | 23.85 | 0.75 - 39.79 | 0.0-0.3 |
| IL-6 (peak; $\text{pg/mL}$) | 189 | 7.7 - 3016 | 0.0-5.0 |
| Absolute lymph count (low; $\text{K}/\mu\text{L}$) | 0.4 | 0.2 - 3.8 | 0.9 – 3.2 |
| Platelet count (low; $\text{K}/\mu\text{L}$) | 140 | 47 - 329 | 160 - 400 |

\*F = female, M = male

**Supplementary Table 5.** COVID-19 immunoglobulins in matched plasma and cerebrospinal fluid at the time of lumbar puncture.

| **Immunoglobulin** | **Plasma** |  | **CSF** |  |
| --- | --- | --- | --- | --- |
|  | **n** | **%** | **n** | **%** |
| Anti-N Protein IgG (Quantitative) | 10/12 | 83.3 | 1/12 | 8.3 |
| Anti-S Protein IgG (Quantitative) | 10/12 | 83.3 | 0/12 | 0 |
| Anti-N Protein IgM (Qualitative) | 4/9 | 44.4 | 0/9 | 0 |
| Anti-N Protein IgA (Qualitative) | 6/9 | 66.7 | 0/9 | 0 |

**Supplementary Table 6.** Demographics and comorbidities of control and COVID-19 CSF patient cohorts.

|  |  | **CoV- cohort, n = 10** |  | **CoV+ cohort, n = 10** |  |
| --- | --- | --- | --- | --- | --- |
|  |  | n or value | Range or % | n or value | Range or % |
| **Age** (mean) |  | 69 | 58 - 77 | 64 | 47 - 79 |
|  | ≥ 65 | 8/10 | 80.0% | 5/10 | 50.0% |
|  | < 65 | 2/10 | 20.0% | 5/10 | 50.0% |
| **Sex** |  |  |  |  |  |
|  | Female | 8/10 | 80.0% | 6/10 | 60% |
|  | Male | 2/10 | 20.0% | 4/10 | 40% |
| **Active Primary & Metastatic Cancer** |  | 10/10 | 100.0% | 10/10 | 100.0% |
|  | Lung | 4/10 | 40.0% | 4/10 | 40.0% |
|  | Lymphoma/Leukemia | 2/10 | 20.0% | 2/10 | 20.0% |
|  | Pancreatic | 2/10 | 10.0% | 1/10 | 10.0% |
|  | Breast | 1/10 | 10.0% | 1/10 | 10.0% |
|  | Ovarian | 1/10 | 10.0% | 1/10 | 10.0% |
|  | Thyroid | 0/10 | 0.0% | 1/10 | 10.0% |
| Brain metastases |  | 5/10 | 50.0% | 5/10 | 50.0% |
| Prior CNS-directed radiation |  | 4/10 | 40.0% | 5/10 | 50.0% |
| Cancer treatment within 30 days |  | 7/10 | 70.0% | 8/10 | 80.0% |

**Supplementary Table 7.** MRI parameters used in this study.

| **Field Strength** | **Sequence** | **TR (msec)** | **TE (msec)** | **Flip Angle** | **FOV (mm)** | **Slice Thickness** | **Matrix (frequency x phase)** |
| --- | --- | --- | --- | --- | --- | --- | --- |
| **3 T** | T1W FSE | 2000 | 26 | 111 | 240 | 5 | 320 x 256 |
|  | T2W FSE | 2500 | 120 | 111 | 240 | 5 | 320 x 320 |
|  | T2 FLAIR | 9000 | 120 | 111 | 240 | 5 | 320 x 224 |
|  | SWAN | Minimum | 24.3 | 15 | 240 | 2 | 320 x 224 |
|  | DWI EPI | 8000 | Minimum | 90 | 240 | 5 | 128 x 128 |
| **1.5 T** | T1W FSE | 742 | 11.7 | 160 | 240 | 5 | 256 x 224 |
|  | T2W FSE | 3268 | 102 | 160 | 240 | 5 | 320 x 256 |
|  | T2 FLAIR | 8000 | 120 | 160 | 240 | 5 | 256 x 224 |
|  | SWAN | Minimum | 50 | 30 | 240 | 3 | 256 x 256 |
|  | DWI EPI | 8000 | Minimum | 90 | 240 | 5 | 128 x 128 |

